# Defining Facets of Social Distancing during the COVID-19 Pandemic: Twitter Analysis

**DOI:** 10.1101/2020.04.26.20080937

**Authors:** Jiye Kwon, Connor Grady, Josemari T. Feliciano, Samah J. Fodeh

## Abstract

Social distancing has been one of the primary mitigation strategies in the United States to control the spread of novel coronavirus disease (COVID-19) and can be viewed as a multi-faceted public health measure. Using Twitter data, we aim to (1) define and quantify the prevalence and evolution of facets of social distancing during the COVID-19 pandemic in the US in a spatiotemporal context and (2) examine the most amplified tweets among social distancing facets. We analyzed a total of 259,529 unique tweets containing “coronavirus” from 115,485 unique users between January 23, 2020 and March 24, 2020 that were identified by the Twitter API as English and U.S.-based. Tweets containing specified keywords (determined *a priori*) were grouped into six social distancing facets: implementation, purpose, social disruption, adaptation, positive emotions, and negative emotions. Tweets about social disruptiveness were most retweeted, and implementation tweets were most favorited. Social distancing tweets became overall more prevalent in the U.S. from late January to March but were not geographically uniform. In January and February, facets of social distancing appeared in Los Angeles, San Francisco, and Seattle, which were among the first cities impacted by the COVID-19 outbreak. Tweets related to the “implementation” and “negative emotions” facets of social distancing largely dominated in combination with topics of “social disruption” and “adaptation”, albeit to a lesser degree. Social distancing can be defined in terms of facets that respond and represent certain moments and events in a pandemic, including travel restrictions and rising COVID-19 case counts. For example, in February, Miami, FL had a low volume of social distancing tweets but grew in March which corresponded with the rise of COVID-19 cases in the city. This suggests that overall volume of social distancing tweets can reflect the relative case count in respective locations.

## INTRODUCTION

The emergence of novel coronavirus disease (COVID-19) and its etiologic cause, severe acute respiratory syndrome coronavirus 2 (SARS-CoV-2), has prompted an international effort to limit its morbidity and mortality. Social distancing has been one of the primary mitigation strategies in the United States, which compels individuals to stay at home as much as possible and avoid close contact with others to reduce transmission and intensity of the pandemic[1-3]. The realization of social distancing and its intersection with many aspects of our social, educational, professional, and emotional lives is unprecedented in its degree of magnitude. Social distancing can be viewed as a multi-faceted public health measure, involving many stakeholders, practices, and consequences. Development and evaluation of a measure of communication on a public health intervention—here, social distancing—may be beneficial for public health and public policy officials [4].

We define social distancing as a multi-faceted intervention where these facets or stages unfold as we are living through the COVID-19 experience. These facets may change in our subsequent studies as COVID-19 events evolve. The facets of social distancing are: (1) *<u>purpose and justification</u>* of imposing this disruptive nation-wide behavioral measure. The purpose is to slow the spread of COVID-19 to levels manageable by healthcare systems. (2) *<u>Implementation</u>* of social distancing to not only avoid mass gatherings but also maintain a 6-feet distance amongst individuals. The advisories imposing this facet translated to closing non-essential businesses, restaurants, schools, and colleges. (3) *<u>Social activity disruptions</u>* which impose travel restrictions and emphasize less human face-to-face interactions; an abrupt change in individuals’ and communities’ highly interconnected networks. (4) *<u>Adaptation</u>* to social distancing by accepting a new way of life and virtually conducting daily life activities. Examples include online schooling, working remotely through teleconferencing, online food shopping, telehealth-based visits as well as online entertaining through platforms such as Netflix. (5) *<u>Positive emotions</u>* and (6) *<u>negative emotions</u>* facets associated with the emotional response to social distancing. These facets could potentially measure the levels of distress culminating over time as a result of disrupting social behaviors and activities that are usually associated with mental and emotional wellness.

Twitter is a microblogging platform by which users (tweeters) socialize and tweet through the network. Users on Twitter can contribute original content through tweeting thoughts, opinions, and news, and engage with existing content by retweeting, favoriting, and replying to others’ tweets. Non-protected tweets are publicly available and approximately 3% of tweets are voluntarily geotagged[5]. As Twitter has the advantage of real-time content availability, it has been harnessed during past infectious disease outbreaks to understand networks, gauge public knowledge, and forecast spread [6-12]. In recent studies, we have utilized Twitter to better understand critical social and behavioral outcomes such as suicide risk [13 14]. Our findings have shown that certain patterns of word use on Twitter can define many social behaviors associated with suicide risk. Given that social distancing is the chief social behavioral measure taken to flatten the curve of COVID-19 spread, this study aims to quantify its perception, implementation, and impact on the nation, through Twitter.

In this study, we adopt a supply-based infodemiology approach[4] to analyze facets of social distancing on Twitter with the overarching goal of informing public health policy and practice. Our newly developed information supply measure is the prevalence of tweets concerning the different social distancing facets. We achieve our goal by collecting tweets related to “coronavirus”. We analyze the contents of the tweets and map them into categories that correspond to the relevant social distancing facets. We then quantify the prevalence of these facets across different states and over time. The objective of this study is two-fold: (1) define and quantify the prevalence and evolution of social distancing facets in a US spatiotemporal context and (2) examine the most amplified tweets among social distancing facets.

## MATERIALS AND METHODS

### Data Collection

We downloaded tweets that contained the term “coronavirus” between January 23^rd^ and March 24^th^, 2020 using the Twitter API. For each tweet we had the following variables: the users’ handle, time of tweet, number of retweets and favorited at the time of collection, geographic coordinates, and text of tweet. Only unique tweets written in English and geotagged to the United States were included in the analyses. Our data is missing tweets from February 29^th^ through March 5^th^ and March 10^th^-March 13^th^, 2020. Despite missingness in the data, we were able to capture the overall trends of social distancing facets, thus we skipped imputation. All analyses were performed using R (version 3.6.1), tidytext package (version 0.2.2), and Tableau (version 2020.1.0).

### Identifying and Grouping Tweets into Social Distancing Facets

Keywords pertaining to facets/topics of social distancing were determined *a priori* and were used to collate tweets into six facets (see Appendix 1). Tweets including terms like “flatten the curve” that tell the motivation for social distancing as to limit viral transmission and protect vulnerable populations were included in the “purpose” facet. The “implementation” facet comprised of tweets capturing content related to institutional closures and public health advisories as to limit exposure to others. The “social disruption” group included tweets concerning the cancellation of social events such as parties, mass gatherings, and other disruptions to daily life. The “adaptation” facet is captured by how individuals adapted their livelihood into virtual settings in the form of online social activity, working remotely, and studying remotely. Tweets that contain words like Zoom, teleconferencing, and Netflix were included. Finally, the last two groups, “positive emotions” and “negative emotions” were designed to capture tweets which provided insights into users’ feelings and attitudes related to and coinciding with the COVID-19 pandemic. Facets were not mutually exclusive as a tweet could be assigned to more than one topic.

### U.S. Trends of Social Distancing Facets

The primary analysis concerned a description of the trends of social distancing facets for the entire dataset of U.S. tweets on a daily and weekly basis. The proportion of a given social distancing facet per day is calculated by dividing the number of tweets belonging to that facet by the total number of tweets that day. These proportions were used for relative comparison over the entire study period and compared to events on the ground. The proportion of a given social distancing facet per week is calculated by dividing the number of tweets belonging to that facet by the total number of tweets that week. Trends of these facets were followed, and χ^2^ tests were used to evaluate weekly change in tweet proportions.

### Spatiotemporal Analysis

Geographic coordinates of the tweets from the different social distancing facets were plotted as pie graphs on a map of the US for January, February, and March. The diameter of the pie graphs corresponded to tweet volume in a given month. For simplified visual interpretation, we introduced a month-specific threshold on the number of tweets to be displayed on the map to eliminate noise that does not rise to a meaningful pattern. Hence, only facets that meet the threshold were plotted at corresponding locations.

### Amplified Tweets in Social Distancing Facets

To estimate the relative amplification of social distancing facets on Twitter, we calculated two measures of tweet amplification for each facet: average number of retweets per tweet and average number of favorites per tweet. For retweets in a given facet, we divided the number of retweets of all tweets in a facet by the total number of tweets in that facet. Similarly, we divided the number of all favorites of tweets in a facet by the total number of tweets in that facet to yield a score of average favorites per tweet.

## RESULTS

### Descriptive Analysis

Our final tweet dataset of 259,529 unique tweets from 115,485 unique users demonstrated an increasing level of coronavirus-related content over the study period. The total tweet count grew from 11,240 (4.3%) tweets in the final nine days of January to 33,713 (13.0%) tweets in February, and to 214,576 (82.7%) tweets in March. The number of unique users increased in a similar fashion, from 8,232 in January tweets, and 18,591 in February, to 100,979 in March. The cities with the highest number of tweets changed by month, but in March alone, 8,045 (3.7%) tweets originated from Los Angeles, CA, 6,325 (2.9%) tweets from Manhattan, NY, and 4,145 (1.9%) tweets from Washington, DC.

Themes among the most frequently used hashtags exhibited heterogeneity by month with January tweets using hashtags focused on the early, localized impact of the virus in China, such as #china, #coronavirusoutbreak, and #wuhan. Similar hashtags were observed in February with more pronounced presence; for example, #coronavirusoutbreak increased by 180%. March hashtags overall focused on the U.S. and social distancing with hashtags such as #socialdistancing, #quarantine, and #stayhome. A full list of the top 10 hashtags per month are included in Supplementary Table 2.

### U.S. Trends of Social Distancing Facets

The daily proportions of tweets belonging to the six social distancing facets are shown in Figure 1. In January, during the earlier phase of the COVID-19 outbreak, tweets among the facet “negative emotions” predominated with all other facets relatively less pronounced. “Implementation” grew in February, accounting for about 10% of coronavirus-related tweets, and then exhibited a strong upward trend in March increasing 245% from February 28^th^ to March 23^rd^. “Implementation” tweets comprised 24% of coronavirus-related tweets at their peak on March 23^rd^. Content relevant to “adaptation” such as working from home and studying online in addition to tweets related to the “purpose” of social distancing also increased over the study period, albeit to a lesser degree. “Adaptation” tweets peaked on March 17^th^, accounting for 4.3% of tweets and “purpose” topic tweets peaked on March 24^th^ at 3.1%. Alternatively, tweets concerning “social disruption” peaked on February 3^rd^ with an 88% increase compared to the previous day making up 8.7% of the total tweets. This facet declined steadily before peaking again on February 23^rd^ comprising 6.3% of the total tweets, which is a 122% increase in comparison to the previous day. After March 18^th^, “social disruption” accounted for the lowest number of tweets among the facets. We analyzed the relationship between time (in weeks) and each facet of the social distancing using chi-squared (**χ^2^**) tests. These tests showed that the increase in the proportion of all facets, besides “social disruption”, were statistically significant (*P<.001*), and the decrease in social disruption tweets was statistically significant (*P<.001*).

**Figure 1:**
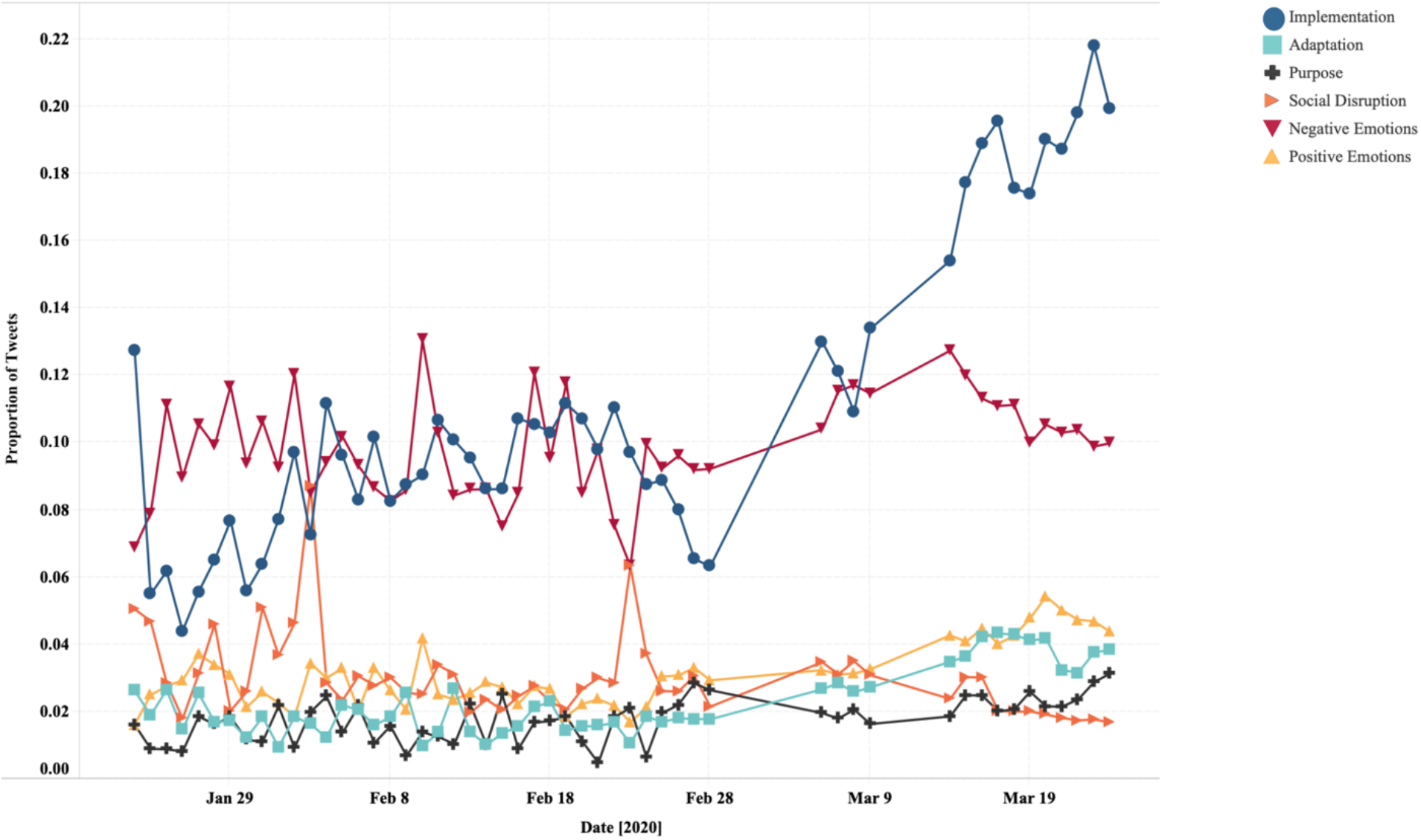
Trends in daily tweet proportions of social distancing facets from January 23^rd^ – March 24^th^, 2020.

### Spatiotemporal Analysis

Social distancing facets were analyzed spatiotemporally using tweets’ geotagged location for January, February, and March (Figure 2 - 4). Each pie graph corresponds to a cluster of tweets such as in a large city or metropolitan area with the diameter being a function of tweet volume, and overlapping graphs simply indicate nearby cities (e.g. Manhattan and Long Island). Over this period, tweets increased in both volume and locations. Areas generating a high number of social distancing tweets included Los Angeles and San Francisco, CA, Manhattan, NY, Washington, D.C., Chicago, IL, Houston, TX, and Tampa, FL.

**Figure 2:**
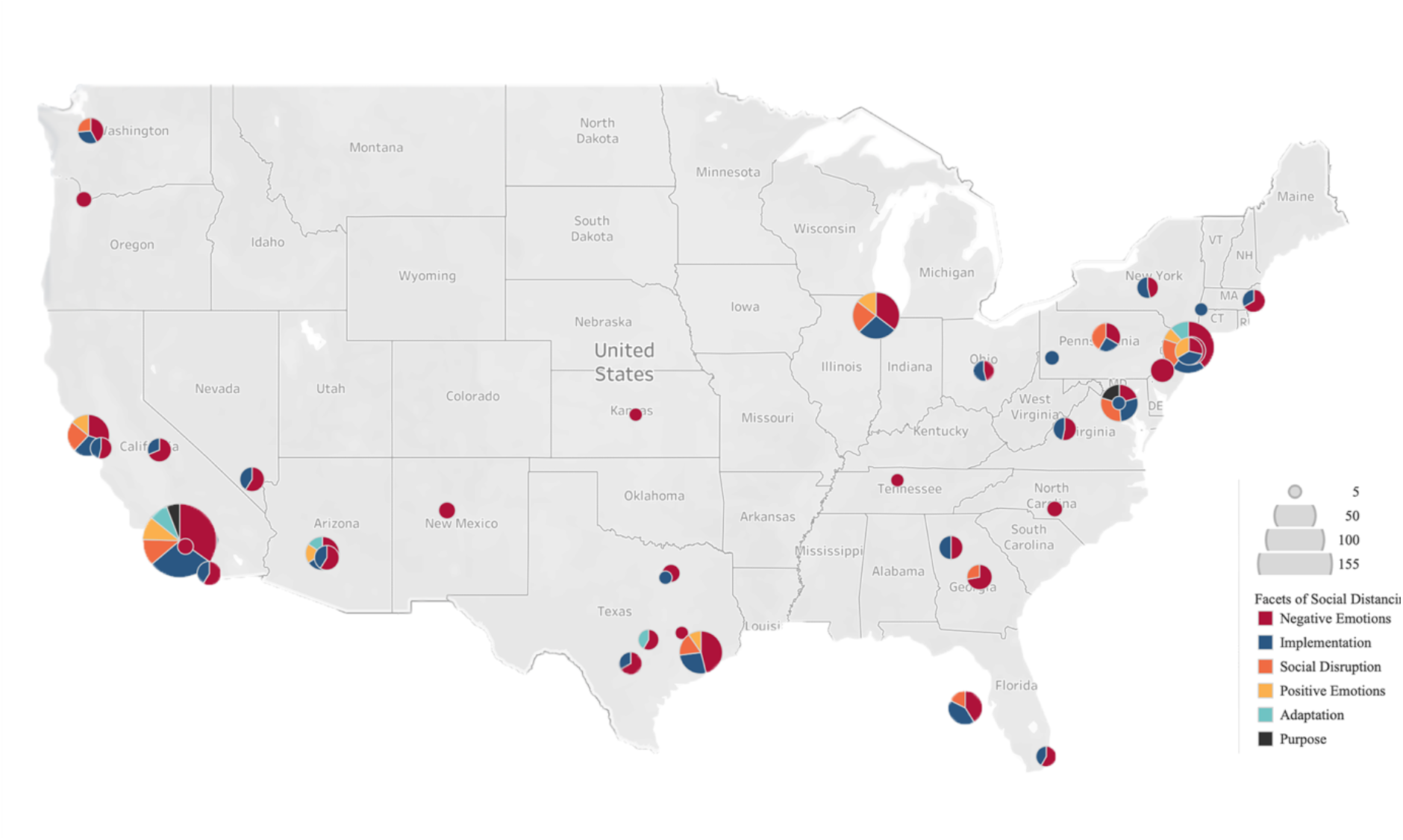
All facets of social distancing mapped for January 2020. Larger diameter denotes higher volume of tweets. Threshold = 5.

**Figure 3:**
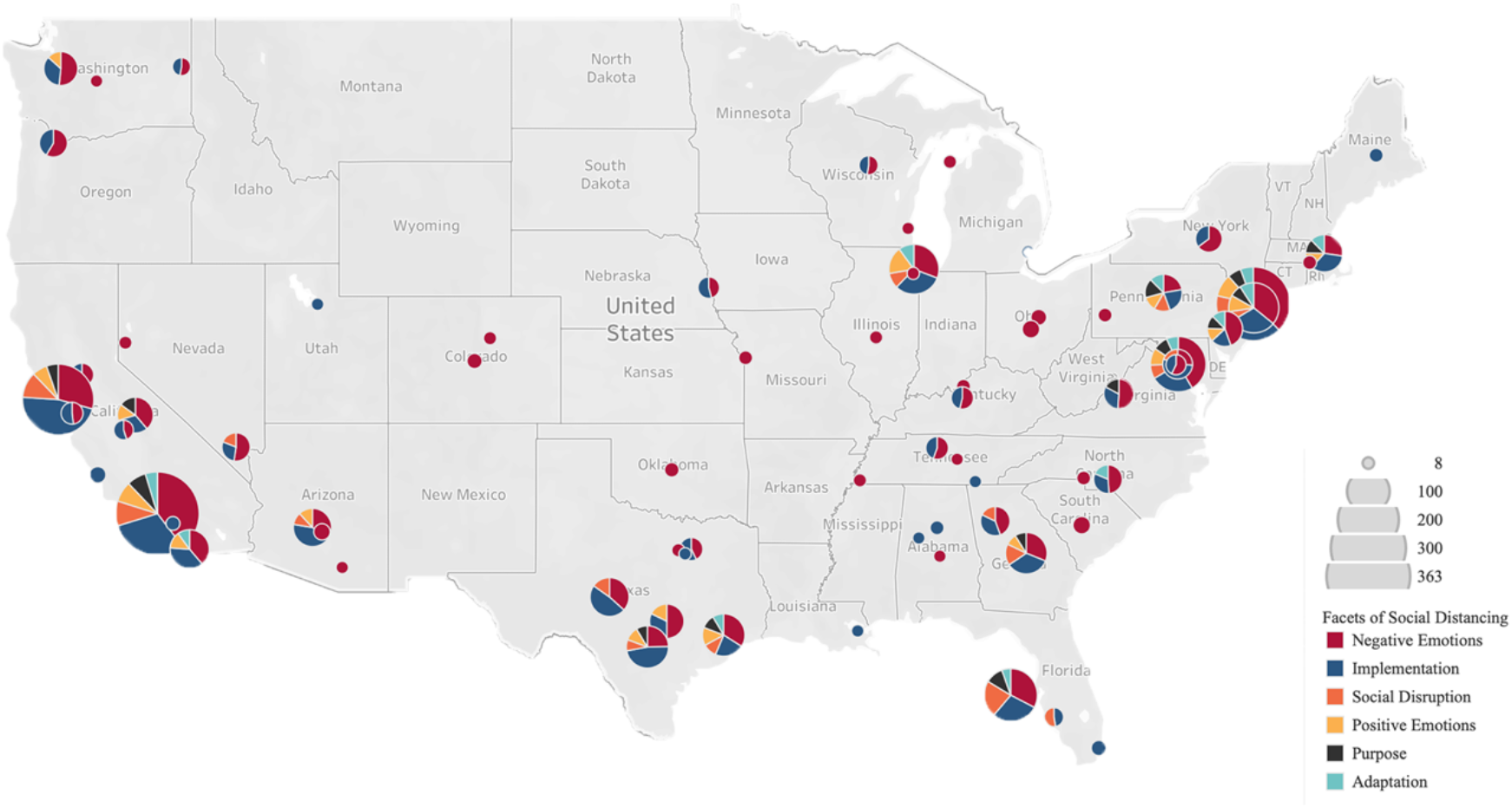
All facets of social distancing mapped for February 2020. Larger diameter denotes higher volume of tweets. Threshold = 8.

**Figure 4:**
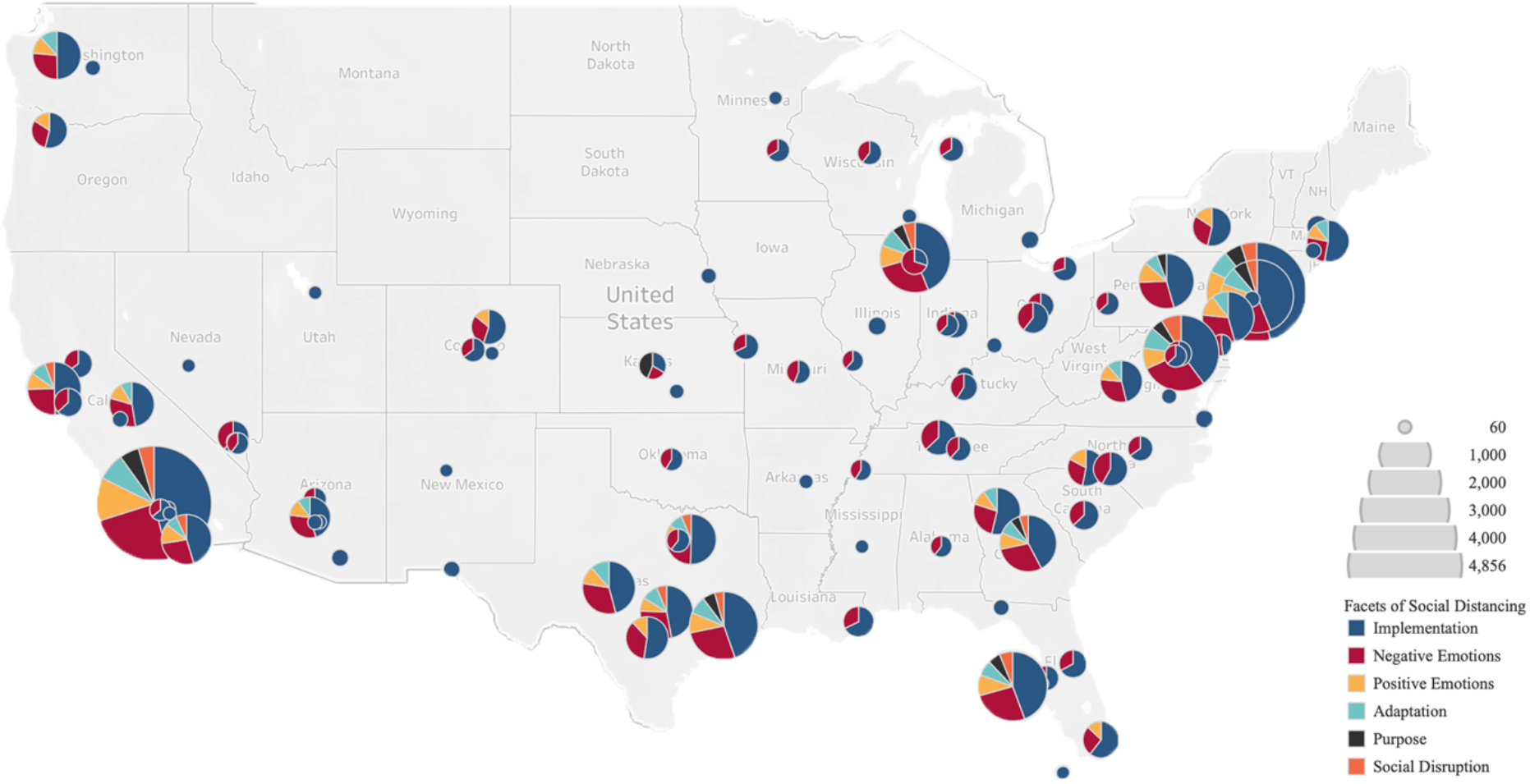
All facets of social distancing mapped for March 2020. Larger diameter denotes higher volume of tweets. Threshold = 60.

The dominant facet per location demonstrates a progression of topic usage. In January and February (Figure 2 and 3), social distancing tweets were sparsely distributed across the states and mostly generated from highly populated areas on the east and west coasts. “Negative emotions” was the most prevalent facet across the states overall, followed by “implementation” and “social disruption”; however, some heterogeneity was observed. Notably, in January, “implementation” was more prevalent in west coast cities such as Los Angeles and San Francisco compared to other metropolitan areas (e.g. New York City) and some cities including Washington, D.C. had a larger relative number of “social disruption” tweets. In February, as the COVID-19 outbreak began to spread internationally, tweets of social distancing facets were generated from more cities (e.g. New York City, Chicago, and Houston) including “implementation”, “social disruption”, “negative emotions”, and “adaptation”.

Figure 4 depicts the growth of the social distancing facets in March 2020 as seen in the large and more numerous pie graphs. Overall, the predominant facet was “implementation”, followed by “negative emotions”. “Adaptation” came next in California, New York, Washington, D.C., Texas, Florida, and Illinois. These observations reflect the onset and enforcement of social distancing on the ground. “Positive emotions” was also pronounced in March mirroring discussions about expectations of social distancing to slow the spread of COVID-19. Tweets relating to social distancing’s “purpose” became more apparent in March, notably in Kansas, Los Angeles, and New York. The large cluster of tweets seen off the coast of western Florida is believed to be a minor fault of the mapping software which plotted tweets from Tampa, FL off the Florida coast. The other tweet coordinates and their plotted locations are taken in confidence as they appear in cities largely impacted by the pandemic.

### Amplified Tweets in Social Distancing Facets

Examining the average number of retweets and favorites of social distancing facets tweets, we noticed that “social disruption” tweets were most amplified through retweeting with 3.74 retweets per tweet on average. This was then followed by “implementation” (3.36 retweets per tweet), “purpose” (3.30 retweets per tweet), “negative emotions” (3.03 retweets per tweet), “positive emotions” (2.22 retweets per tweet), and “adaptation” (2.16 retweets per tweet). On the other hand, “implementation” exhibited the highest number of favorites, an average of 14.84 favorites per tweet, followed by “negative emotions” (12.03 favorites per tweet), “social disruption” (9.7 favorites per tweet), “purpose” (9.4 favorites per tweet), “positive emotions” (9.33 favorites per tweet), and finally, “adaptation” (8.33 favorites per tweet).

## DISCUSSION

In this Twitter analysis of social distancing-related tweets during the COVID-19 pandemic, several observations emerged. During the early phases of the COVID-19 pandemic in January and February, outbreaks were confined to China and nearby countries; consequently, tweets during these early months were thought to be confined and in reference to the situations in these countries. The U.S. saw a dramatic increase in COVID-19 cases in March, prompting intense national social distancing efforts; accordingly, tweets were regarded as referring to U.S. events, attitudes, and reactions.

In January and February, it was shown in Figures 2 and 3 that the prevalence of tweets captured locations that started voicing out through the defined facets. Interestingly, locations captured during the early phases of the outbreak are states that were thought to experience the COVID-19 earlier than other states, including Washington, Illinois, and California. In these two maps, the higher prevalence of the “implementation” and “social disruption” facets in the west coast cities (Los Angeles, San Francisco, and Seattle) compared to other major cities could be attributed to their status as major hubs for international flights, many of which originate from East Asia. The “social disruption” facet was particularly pronounced during the same time in these cities and others including Manhattan, NY and Washington, D.C., albeit more so in January. In February, we observed two notable peaks of “social disruption” (Figure 1) on February 3^rd^ and 23^rd^ that correspond and may be reactionary to travel restrictions taken into effect on the evening of February 2^nd^ and Italy going on a nationwide lockdown on the 23^rd^[15 16]. This facet captured reactions to the imposed advisories and restrictions as it was formed based on words like “travel”. Negative emotions were also highly prevalent in many locations, suggesting an expected reaction to a novel infectious disease of concern, uncertainty, and fear[17-19] among U.S. users while the outbreak was largely confined to China.

Turning to the month of March where social distancing efforts were realized in the U.S. and the number of COVID-19 cases increased dramatically, there were certainly more locations across the country that voiced out about these facets of social distancing. Trends in these facets (Figure 1) demonstrate that tweets relating to the implementation of social distancing increased markedly in March and expanded to many locations across the U.S.. This was likely reactionary to and coincided with nationwide social distancing initiatives such as the Centers for Disease Control and Prevention’s recommendation to cancel events with more than 50 people on March 15^th^ [20] and events with more than 10 people for higher risk populations [21]. The peak of implementation in our study was on March 23^rd^ with 24% of all tweets in our dataset related to social distancing implementation. This was around the same time when New York City was declared the U.S. COVID-19 epicenter[22], and taken together with the large volume of tweets from the New York area in March (Figure 4), perhaps the situation in New York was in part responsible for this peak. Along with rise of implementation, negative emotions increased in early March but subsequentially decreased until March 24^th^ possibly as a result of increased emergence of other facets. Less represented facets, such as “positive emotions”, “purpose”, and “adaptation” were still thought to play key roles in users’ reactions and reflection on social distancing especially across locations such as Los Angeles, TX, FL, CO, and NY. In these locations, pie graphs with larger diameters and higher representations of social distancing facets were seen. The facets “positive emotions” and “purpose” became more pronounced in March, and adaptation reached and maintained its peak across all weekdays on the third week of March as people returned to work and school but on online platforms. This increase in “adaptation” could also explain the decrease in “social disruption”, observed in Figure 1, perhaps as individuals acclimated to the new routines and practices.

Social distancing tweets as a whole were predominantly generated from the Northeast, South, and West. Relating this to the observed case counts on the ground[23], among the list of states with the highest recorded COVID-19 case count included areas in the Northeast, South region and West coast, specifically California. These figures not only reveal locations with high numbers of facets of social distancing tweets but also reveal locations with relatively low tweet volume. For example, in February, Miami, FL has a low volume of social distancing tweets but grew in March which corresponded with the rise of COVID-19 cases in the city[23]. This suggests that overall volume of social distancing tweets can reflect the relative case count in respective locations.

Our secondary research objective examined amplified tweets to further understand the drivers of the public’s perception during the COVID-19 pandemic and a period of intense social distancing. We defined amplified facets as facets which had high relative engagement, measured by an average number of retweets and favorites. As described in the Risk Amplification through Media Spread model, amplified tweets play a key role in the public’s perception and response [24] and signal which content is meaningful to users [25]. Our study showed that the most amplified facet of social distancing tweets measured by the retweet count was “social disruption” and least amplified was the “adaptation” group. In terms of favorites, “implementation” had the highest number of mean favorites per tweet and the least amplified was “adaptation”. These results suggest that not only do users find these topics meaningful and worth engaging with, but also demonstrate that the topics of “implementation” and “social disruption” were highly broadcasted among social networks. As such, these facets can be leveraged to promote public health actions by echoing the wavelength that the public shares.

Our study responds to the growing interest in the application of infodemiology in public health [4]. We define facets of social distancing with the advantages of real-time, publicly available Twitter data. Through infoveillance and infodemiology, previous studies have shown that Twitter may have the potential to serve as an aid for infectious diseases surveillance tool[25]. A longitudinal study like this one is especially useful during an outbreak [4] for informing intervention efforts by providing a closer look at the prevalence of multiple facets of social distancing. Observing the change in social distancing facets mapped through time provides insight into location-specific content, including possibly when certain localities experience cases. Spatiotemporal analysis of tweets may be of higher importance than just temporal analysis which is often performed. Some have argued that temporal analysis coupled with a spatial dimension tend to match the actual infectious disease epidemiology and have potential to detect possible outbreaks or early signals of a potential outbreak [26].

### Limitations

There are several limitations to the current study. Only tweets including the word “coronavirus” were downloaded from the Twitter API and included in the analysis. Over the course of the pandemic, terminology has shifted toward other nomenclature such as COVID-19, SARS-CoV-2, or referred to colloquially as “corona”, and in some circles as the “Wuhan virus” or “China virus”; these tweets were not captured. However, we demonstrated that “coronavirus” was highly used as we collected over 250,000 unique tweets, and this term is the most consistently used term to describe the crisis as this name preceded others. Additionally, the number a tweet has been retweeted is dependent on when the data is collected. Our data collection practices were not consistent in regard to time of day. Nevertheless, given the long period of data collection, this should not be concerning. Finally, tweets belonging to positive and negative emotion facets were classified in a way that did not necessitate they be in regard to social distancing topics (as did other facets) but only to coronavirus. Still, these tweets are useful as they coincide with intensive social distancing efforts and thus offer important insight into how individuals reacted emotionally during this period.

## CONCLUSION

We conclude that social distancing can be defined in terms of facets which may respond to certain moments and events in a pandemic. Social distancing efforts during the COVID-19 pandemic are unprecedent and measurement of these practices is challenging, but Twitter can be applied to understanding the public’s practice of and response to social distancing. The spatiotemporal analysis of multiple facets of social distancing in this study helps evaluate the penetration of information and has the potential to provide insights in evaluating public health measures.

## Data Availability

Publicly available Tweets downloaded using the Twitter API.

## ACKNOWLEDGEMENTS

The authors would like to thank Dr. Daniel Weinberger for his contributions to the early stages of this project and Dr. Glen Nowak for his inputs on Twitter methodology and guidance on health communication.

## CONTRIBUTOR STATEMENT

JK and CG contributed to the concept, analysis and interpretation of results, acquisition of data and drafting/revising the manuscript. JF contributed to the acquisition of data, implementation of analytical approaches and reviewing the manuscript. SF contributed to concept and design, analysis and interpretation of data, drafting/revising the manuscript, critical revisions of the manuscript for important intellectual content, and supervision including responsibility for the conduct and final approval.

**Appendix 1.**
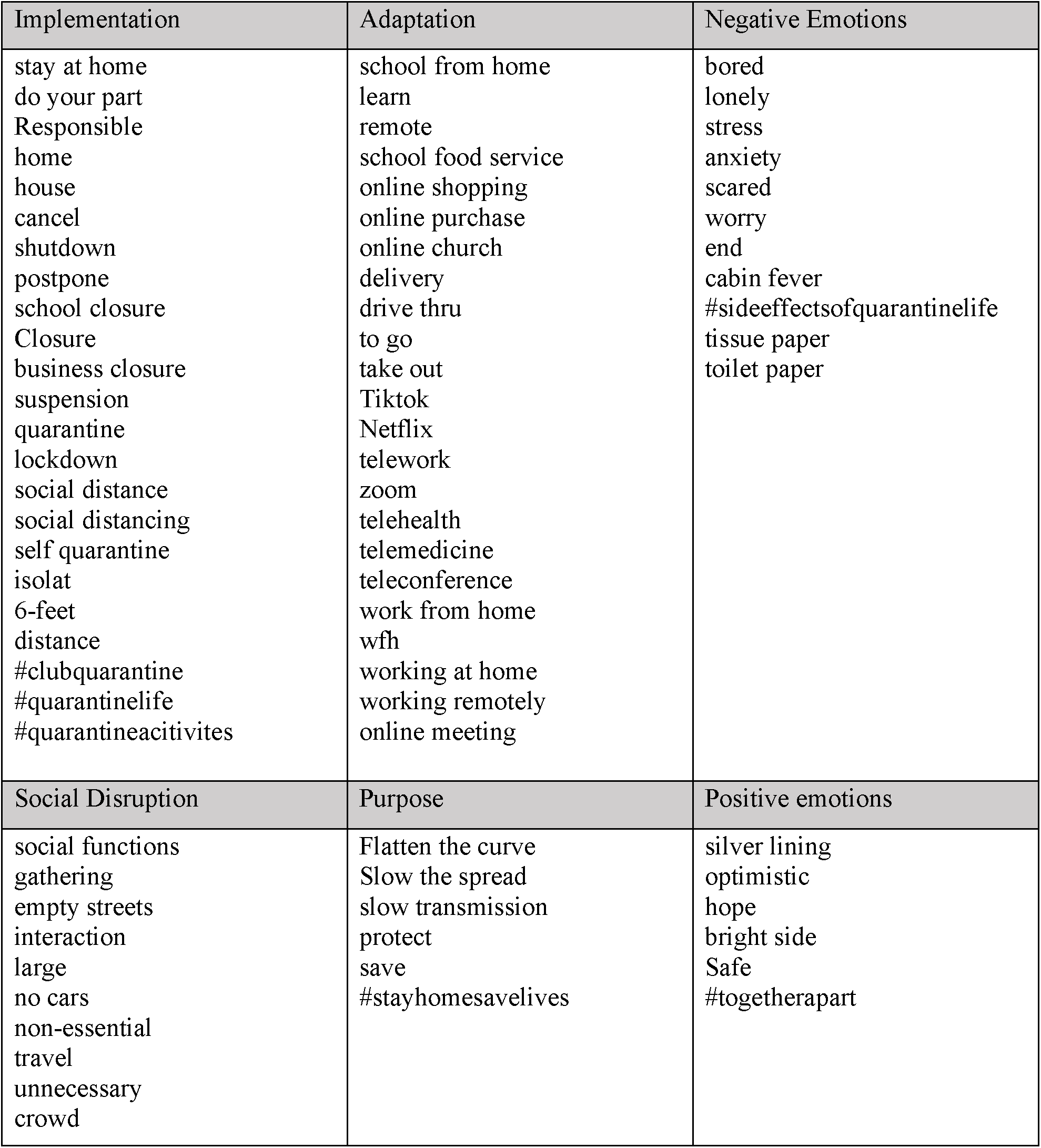
Keywords for each facet of social distancing

**Appendix 2.**
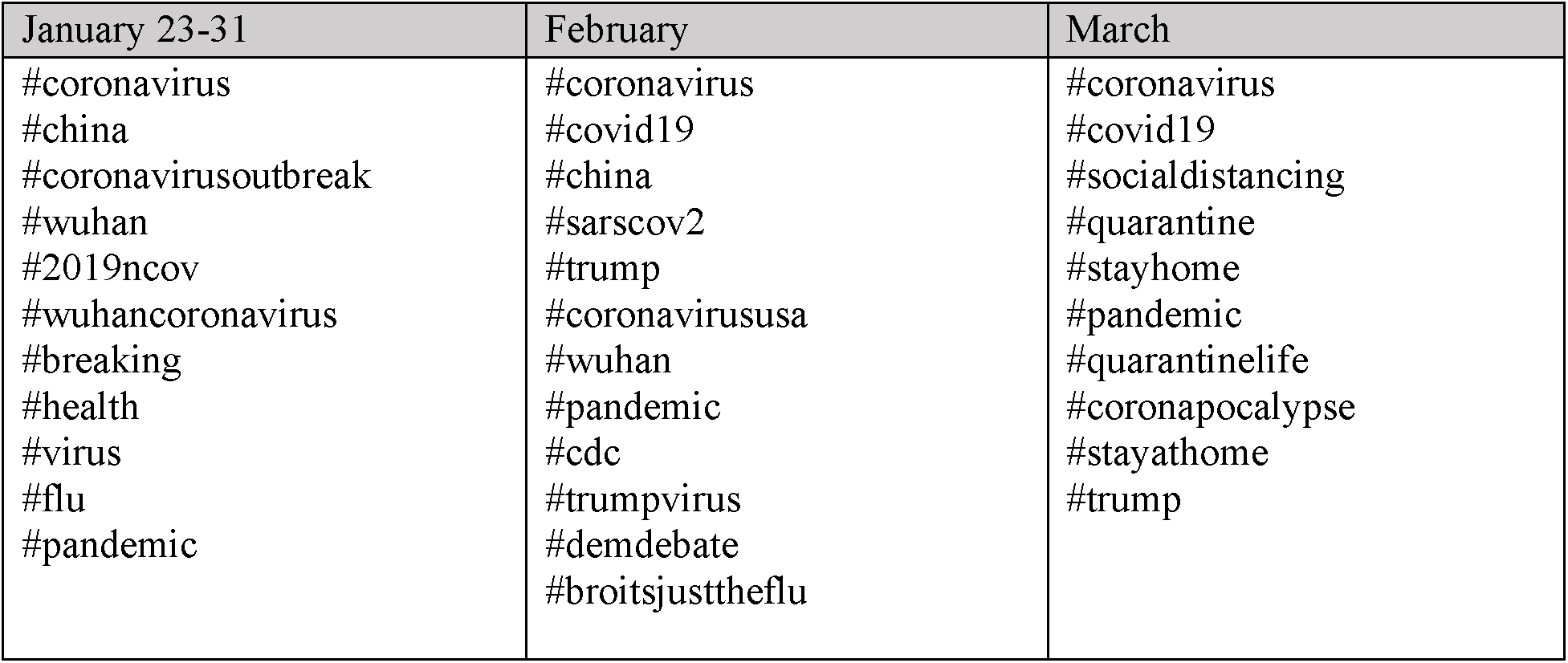
Most frequently used hashtags by month

